# A Growing Burden, a Persistent Gap: Cannabinoid Hyperemesis Syndrome Education and Emergency Department Trends in Michigan’s Legal Cannabis Market

**DOI:** 10.64898/2026.10.09.26365113

**Authors:** Elizabeth R. Lorbeer, Nicholas Bronson, Luke A. Burgard, Mary E. Engle, Devjit S. Randhawa, Phoebe V. Sotiroff, Hannah R. Walker, Hanglin Zhu, Joshua D. Mastenbrook

## Abstract

**Background:** Since Michigan’s legalization of recreational adult-use cannabis in December 2018, health care providers in the city of Kalamazoo, Michigan, have informally observed an increase in cannabinoid hyperemesis syndrome (CHS) presentations, although the extent of this increase has not been formally characterized locally. CHS is frequently underrecognized and misdiagnosed, often resulting in repeated emergency department (ED) visits and substantial diagnostic costs before a diagnosis is reached. Despite this clinical and financial burden, it is unclear whether consumers are adequately informed of this risk through point-of-sale or public health resources.

**Methods:** A cross-sectional environmental scan was conducted to identify publicly available educational content about CHS on the websites of licensed cannabis retailers in Kalamazoo and relevant public health agencies. Provisioning centers and public health websites were screened using a structured coding rubric developed specifically for the environmental scan to assess the presence, depth, web page placement, and intended audience of CHS-related content. Additionally, the electronic medical records at a local teaching hospital in Kalamazoo were queried to identify ED visits with a primary diagnosis of CHS from January 1, 2010 to December 31, 2025.

**Results:** Before the legalization of marijuana, the hospital recorded six ED encounters with the primary diagnosis of CHS between January 2010 and November 2018, and 274 ED encounters between December 2018 and December 2025 (mean age: 29 years; 157 females, 123 males). The incidence rate of CHS-related ED encounters was 57.2 times higher during the post-legalization period than during the pre-legalization period (95% CI, 26.0-157.3; exact Poisson rate-ratio test, p < 0.0001). Among the licensed Kalamazoo provisioning centers reviewed, only one mentioned CHS as a warning but did not describe symptoms or offer medical guidance. CHS content was not identified on the websites of the state or local health departments.

**Conclusion:** Despite the growing clinical concern regarding CHS, consumer-facing educational resources remain scarce. Although this study cannot establish a causal relationship between the availability of CHS-related educational content and the observed increase in ED encounters following legalization, it identifies a substantial lack of CHS-specific warnings in the evaluated consumer-facing resources. Given the clinical and potential economic burden associated with delayed CHS recognition, we recommend that Michigan’s existing point-of-sale pamphlet requirement be amended to include CHS-specific warning language.

## Introduction

Cannabinoid hyperemesis syndrome (CHS) is a chronic, heavy cannabis use condition characterized by recurrent episodes of severe nausea, vomiting, abdominal pain, and compulsive hot bathing behavior. Although CHS was first described in the medical literature in 2004, it remains an underrecognized clinical and public health concern despite the legalization of cannabis.^1^ Understanding whether consumers are being informed about the risk of CHS and whether these warnings are reaching them at the point-of-sale has become an increasingly important public health question, as more states permit the sale of recreational cannabis.

Michigan legalized medical marijuana in 2008 and adult-use cannabis in December 2018, becoming the tenth state to legalize recreational cannabis for adults 21 years old and older.^2,3,4^ Prior to recreational legalization, a CDC-funded study revealed that approximately one in nine Michigan residents (11.6%) reported monthly cannabis use, a prevalence exceeding both regional and national averages and representing a 60% increase between 2002 and 2017.^5^ Since legalization, Michigan has become one of the largest legal cannabis markets in the United States, ranking second in total cannabis sales.^6^ The state does not cap the number of retail licenses for provisioning centers at the state-level. Instead, municipalities decide whether to permit cannabis businesses to operate.

The city of Kalamazoo has a population with approximately 73,000.^7^ It hosts substantial health care and educational infrastructure, including two major teaching hospitals affiliated with Western Michigan University Homer Stryker M.D. School of Medicine and Western Michigan University’s College of Health and Human Services.

Against this backdrop, clinical teachers at the School of Medicine anecdotally began reporting an increase in the number of patients presenting with CHS symptoms. A preliminary review of the electronic medical records for the keywords “cannabinoid hyperemesis syndrome” and “cannabis hyperemesis syndrome” was conducted to better understand the local impact of this issue and identified a notable increase in emergency department (ED) presentations following the legalization of recreational marijuana sales.

Despite this rise in cases, it remains unclear whether public health messaging and educational materials are available. Previous studies have examined the web content of medical cannabis, documented educational material gaps in dispensaries, and described the lack of regulatory warning labels across states.^8,9,10^ However, no prior study has specifically characterized the availability of CHS-focused consumer-facing content across the channels most likely encountered by consumers at retail points of sale and local or state public health websites in a jurisdiction where cannabis sales are permitted. If gaps in CHS educational materials exist within an environment as well-resourced as Kalamazoo, they may signal broader opportunities to improve public awareness and harm-reduction efforts in other communities. To address this gap, our study seeks to answer the following research questions:

1. What is the availability of consumer-facing educational content on CHS or the signs and symptoms of cannabis use disorders on the public websites of licensed provisioning centers?
2. What is the availability of consumer-facing and provider-oriented educational content addressing CHS on local and state public health department websites?
3. What was the local ED burden of CHS at a Kalamazoo-area teaching hospital before and after Michigan’s recreational cannabis legalization?
4. Can a standardized, low-resource environmental scan protocol be developed and piloted for replication by other communities to evaluate the availability of CHS educational resources?

## Methods

A cross-sectional environmental scan was conducted to characterize the current landscape of publicly available consumer-facing educational content on CHS. This study examined consumer-facing CHS information availability in a city with an actively licensed medical and adult-use cannabis market.

The scan comprises of two components: (1) a review of the public-facing websites of licensed cannabis provisioning centers operating in the city and (2) a review of the Kalamazoo County Health and Community Services Department (KCHCSD) website and the Michigan Department of Health and Human Services (MDHHS) website for consumer-facing or health care provider-oriented content addressing CHS, both of which were accessed and reviewed in July 2026.^11,12^ To provide a clinical context for these content-availability findings, we additionally characterized the local ED burden of CHS at a teaching hospital in Kalamazoo.

### Setting

This study was conducted in the city of Kalamazoo, Michigan, where licensed medical and adult-use recreational cannabis provisioning centers are legally permitted to operate. For provisioning centers that operate on a website that serves multiple states, the coder was instructed to review only the web pages for provisioning centers that operate within the city.

### Selection criteria

Licensed cannabis provisioning centers operating in the city’s zip codes were identified using the Michigan Cannabis Regulatory Agency’s (MCRA) public licensee database, which was accessed on July 15, 2026.^13^ If the provisioning center had a MCRA record type of “Marihuana Retailer-License,” both medical and adult-use provisioning center licenses were eligible for inclusion. (The spelling with an “h” of marihuana is used in Michigan law). Provisioning centers were excluded if their license status was void as of the access date. Of the 34 licensed provisioning centers identified as “Marihuana Retailer-License,” 19 met the inclusion criteria.

The KCHCSD and the MDHHS websites were included by design as the relevant county- and state-level public health department sites relevant to this scan.

### Coding procedure

Two reviewers independently coded each website using a researcher-developed structured coding rubric that assessed the presence, depth, web page placement, and intended audience of CHS-related content. The rubric was developed specifically for this environmental scan to provide standardized operational definitions and consistent classification of CHS-related website content. The coding instructions and instrument are provided in the Supplementary Materials. The rubric was piloted on a sample of 10 websites before the full data collection. Following the pilot, the reviewers independently applied the rubric to the full sample size. Discrepancies between the reviewers were resolved through discussion and consensus.

### Data collection

For the Kalamazoo provisioning center websites, two reviewers used the Google search string site:[domain] CHS OR “cannabinoid hyperemesis” OR “cannabis hyperemesis” to locate relevant subpages for each identified domain. The same search strategy was applied systematically across all identified provisioning center websites. For the websites of the county and state health departments, the Google search site:[domain] CHS OR “cannabinoid hyperemesis” OR “cannabis hyperemesis” was used. The “CHS” term was omitted from the county and state health department scans because of the ambiguity of the acronym and its association with multiple unrelated terms. Where relevant subpages were located, the reviewers examined the content in full for CHS-related material and recorded the search string and coding results in the study spreadsheet.

### Data capture and archiving

Given the dynamic nature of website content, each page containing CHS-related educational content was archived using a screenshot and saved as a PDF to preserve verifiable content records. The coded data were recorded and stored in spreadsheets, and the resulting dataset was deposited in the Figshare repository.^14^

### Clinical burden query

We queried the Epic SlicerDicer self-service reporting tool at one of the Kalamazoo-area teaching hospitals for all ED encounters with a primary diagnosis of CHS. We queried two overlapping date ranges: (1) January 1, 2010 to December 31, 2025, to capture the full available record, and (2) December 1, 2018 to December 31, 2025, corresponding to the period following Michigan’s recreational cannabis legalization, to allow comparison of pre- and post-legalization encounter counts.

### Data analysis

ED encounter rates for CHS in the pre-legalization period (01/01/2010 to 30/11/2018) and post-legalization period (01/12/2018 to 31/12/2025) were compared using an exact Poisson rate-ratio test, with person-time as the denominator for each period. Analyses were performed in R version [4.5.3] using the poisson.test() function.

### Ethical considerations

This study received an IRB determination of non-human subjects research, as all the data were provided in an aggregated format and no interaction or intervention with provisioning center staff or patients occurred.

## Results

Between January 1, 2010, and December 31, 2025, the hospital recorded 280 ED encounters with a primary diagnosis of CHS (mean age, 29 years; female = 157 [56.1%], male = 123 [43.9%]). The overwhelming majority of encounters, 274 (97.9%), occurred during the post-legalization period (December 1, 2018 to December 31, 2025), whereas only six encounters (2.1%) occurred during the 8.9-year pre-legalization period (January 2010–November 2018) (Table 1).

**Table 1:** Description of ED visits related to CHS.

| **Metric** | **Pre-Legalization**<br>(01/01/2010 – 30/11/2018) | **Post-Legalization**<br>(01/12/2018 – 31/12/2025) | **Study Period**<br>(01/01/2010-31/12/2025) |
| --- | --- | --- | --- |
| Total duration (years) | 8.9 | 7.1 | 16 |
| Total number of ED encounters | 6 (2.1%) | 274 (97.9%) | 280 (100%) |
| Average Age |  | 29 | 29 |
| Female, n | 5 | 152 | 157 |
| Male, n | 1 | 122 | 123 |
| Encounters per year | 0.67 | 38.59 | 17.5 |

The incidence rate of CHS-related ED encounters was 57.2 times higher in the post-legalization period than in the pre-legalization period (95% CI, 26.0-157.3; exact Poisson rate-ratio test, p < 0.0001).

### Analysis of licensed provisioning centers

Among the 19 Kalamazoo cannabis businesses with active licenses, only one referenced cannabis hyperemesis syndrome (CHS) by name as a potential side effect of cannabis use. This mention was classified as “buried,” as it appeared only within an Education Booklet rather than on the main website, and it did not include information about specific symptoms or guidance on when to seek medical care. No other reference to CHS was found elsewhere on the business’s website. The content was identified using the study team’s search.

### Analysis of Public Health Departments

Upon surveying online health department resources for Kalamazoo County and the State of Michigan, CHS mentions were not found on their respective websites or through our search strategy.

## Discussion

This study revealed a substantial increase in CHS-related ED encounters in Kalamazoo following the legalization of recreational marijuana in Michigan, alongside a marked absence of point-of-sale and public health information about the condition. Although this study cannot establish a causal relationship between cannabis use and CHS, this increased incidence raises two distinct questions: Whether increased cannabis use following legalization is producing more actual cases of CHS, and separately, whether increased clinical awareness is improving the recognition and diagnosis of a condition that was previously underdiagnosed.

### Regulatory background

The Michigan Legislature requires all marijuana sold through provisioning centers to include the following warning on the exterior of the packaging:

Marijuana use by pregnant or breastfeeding women, or by women planning to become pregnant, may result in fetal injury, preterm birth, low birth weight, or developmental problems for the child. ^15^

Currently, the law states that a pamphlet is to be available to every customer at the time of sale, which includes safety information related to marijuana use by minors and contains the Poison Control hotline number. The Kalamazoo City Code goes further, requiring retailers to prominently display a sign with the same warning near the entrance of the business.^16^

Opposition to expanding warnings to consumers is documented in Michigan’s legislative record, in which a House Fiscal Agency analysis of the bills that established this warning noted that some stakeholders opposed further labeling, arguing that requiring warnings on a large number of products already carrying the required disclosures could dilute the overall effectiveness of warnings by making them too common to be noticed.^17^ This concern may explain the continued absence of a CHS-specific warning, despite the clinical burden observed in this study. The absence of such a warning may also reflect retailers’ financial incentives, as messaging about CHS risks could discourage high-frequency purchases among consumers at greater risk of developing CHS. Provisioning centers have little incentive to prominently display warnings that could potentially reduce sales. Consequently, consumers and the public have limited opportunities to learn about this condition before symptoms appear and prompt an ED visit.

### Federal executive order

Recent federal policy developments have added further relevance to local surveillance efforts. In December 2025, Executive Order 14370, *Increasing Medical Marijuana and Cannabidiol Research*, directed federal agencies to expedite the rescheduling of marijuana under the Controlled Substances Act and to develop research methods using observational evidence to assess health outcomes associated with medical marijuana and legal cannabidiol (CBD) products.^18^ Notably, the order explicitly calls for this research to focus on long-term health effects in vulnerable populations, including adolescents and young adults. The order also acknowledges that decades of federal Schedule I status have limited the research infrastructure needed to inform the public about safety and efficacy, leaving a documented evidence gap that this study’s environmental scan approach is well-positioned to fill at the local level. This emphasis on observational evidence aligns well with the low-cost community surveillance of adverse outcomes, particularly as cannabis potency and accessibility continue to increase with legalization.

### Economic burden

The reluctance of retailers to disclose CHS risk effectively shifts the cost of this risk onto the health care system. A patient presenting to the ED with abdominal pain, nausea, and vomiting undergoes a broad diagnostic workup, because the differential diagnosis for this constellation of symptoms is extremely broad. CHS is typically a diagnosis of exclusion, considered only after gastrointestinal, obstetric, infectious, metabolic, toxicologic, and neurologic etiologies have been ruled out. ^19,20^ Physicians should consider the potentially dangerous causes of this clinical presentation before moving on to less threatening possibilities. This can prompt extensive resource utilization and repeated diagnostic evaluations, potentially delaying CHS diagnosis and increasing health care charges. The average cost of a CHS visit has been on the rise since the legalization of cannabis. A retrospective observational cohort study of 458 patients with nausea and vomiting who presented to various clinical sites with suspected CHS incurred significantly higher health care costs than controls did.^21^ This study revealed that patients with CHS had a mean cost of $1,425 USD per ED encounter and $4,095 per hospital admission, with a substantial proportion of these expenditures attributable to radiological imaging as part of the diagnostic workup. Another retrospective cohort study examining CHS hospitalizations in Massachusetts from 2012 to 2021 reported that the mean hospital visit cost was $7,460 before the legalization of cannabis and increased to $18,714 after legalization.^22^ This period was also accompanied by an increase in the length of stay and thus in the use of health care resources. Importantly, we surmise that reported health care expenditures may not fully capture the costs associated with complications of prolonged nausea and vomiting and thus underestimate the broader economic implications of CHS. In addition to direct health care costs, severe CHS symptoms may increase ED utilization and resource demands through repeated diagnostic evaluations.

### Prevention rationale

CHS is becoming a potentially serious public health concern that affects the health of communities and burdens local hospital systems. The increased availability of consumer-facing educational content is uniquely positioned to combat this condition by serving as a form of both primary and tertiary prevention measures. As a primary prevention tool, disclosing the potentially adverse health effects of substances should be a central component of their distribution. Just as it is the ethical duty of clinicians to educate patients on the side-effect profile of the pharmaceuticals they recommend, licensed dispensaries should warn their consumers of the potential health threats caused by marijuana consumption. This is a salient point as our society moves toward increased acceptance of naturopathic or alternative medicine, as it would be advisable to set the precedent now. Tertiary prevention, increased educational content, and research efforts on cannabis topics will increase awareness on both sides of the clinical encounter, expediting the diagnostic process, and improving treatment and outcomes. Increasing the frequency with which cannabis use and its adverse effects appear in marketing media may help normalize this topic in conversation. The aim is to reduce the stigma and increase patients’ willingness to disclose cannabis use to their clinicians. A recent exploratory study found that 21.2% of patients never disclosed their cannabis use to health care providers.^23^ Promoting more transparency at the point-of-sale about disclosing cannabis use in the health care setting may help to alleviate and reduce potential stigma surrounding cannabis use. By facilitating patient disclosure of cannabis use, clinicians may be able to obtain more complete and accurate histories when interviewing patients with severe nausea and vomiting.^24^

These findings may interest policymakers in evaluating current point-of-sale disclosure requirements for cannabis. Because CHS is not currently included in Michigan’s mandated pamphlet content, incorporating CHS-specific information may represent a low-burden regulatory update by the MCRA rather than requiring new legislation. Additionally, local and state public health websites represent an opportunity to improve consumer awareness. Although public health websites provide an accessible venue for consumer education, point-of-sale and product-level messaging may offer a more direct opportunity to reach cannabis consumers with CHS information.

### Limitations

This study was limited to one city in Michigan, yielding a small sample of dispensaries. ED encounters were assessed from January 2010 to December 2025, while retailer and public health websites were reviewed in July 2026. Thus, it is unknown whether CHS-related information was available to consumers throughout the encounter period. Printed pamphlets were not posted on provisioning center or public health websites. Therefore, we could not confirm whether CHS information was present on them. Notably, the single pamphlet we located listed “Cannabinoid hyperemesis syndrome (CHS)” as a potential risk or side effect but provided no CHS-specific description, symptoms, or guidance on what to do or when to seek care.^25^ We also lacked insight into verbal communication between consumers and distributors at the point-of-sale, so it is unclear whether verbal CHS education is occurring, and it is similarly unknown whether patients received oral or printed education on CHS at the time of ED discharge. Because the study design does not support causal inference, these findings are best understood as complementary indicators of local CHS burden and the current state of consumer education, rather than as directly linked exposure and outcome data.

A further limitation concerns the structure of the encounter data itself. This analysis compared aggregate encounter counts across two defined periods (pre- and post-legalization) rather than evaluating within-period trends. Monthly or yearly data, which were unavailable, would have allowed for the characterization of the trajectory of change (e.g., gradual increase vs. abrupt shift). The small number of pre-legalization encounters (n = 6) also produced an imprecise rate estimate, as reflected in the wide 95% CI (26.0–157.3), meaning that a single additional or missing case could meaningfully alter the observed rate-ratio. Additionally, it is unclear whether the low number of documented CHS ED diagnoses in the pre-legalization period was due to fewer cases of CHS, provider unawareness or non-consideration of the condition, or simply coding the final diagnosis as something other than CHS.

### Future directions

During our environmental scan in the data collection period of this study, one business notably removed its existing cannabis educational page from its website. The page was identified only through the Google search query used in the environmental scan, allowing us to retrieve the previously indexed content. The exact motivation for this change remains unclear. This observation illustrates how retailer-specific educational efforts can be dynamic and impermanent, making them likely insufficient to independently address the CHS burden. When left to the discretion of individual businesses, the availability of educational content may vary considerably. Therefore, lawmakers should prioritize the standardization of this process.

Future studies could assess CHS awareness among bud tenders, health care providers, and public health workers to determine whether limited awareness exists on the distribution side, the clinical side, or both. Where gaps are identified, targeted training could support earlier recognition and reduce financial burden and ED visits. Other directions include comparing CHS trends across cities and states that legalized cannabis earlier than Michigan did, such as Colorado, which legalized medical use in 2000 and recreational use in 2012.^26^ Given the additional time since legalization, these states likely have accumulated more CHS cases and may have adopted different requirements for warning consumers. Future studies could also evaluate different formats of educational materials for informing the public about CHS, and monitor whether their distribution corresponds with changes in local CHS-related ED trends. Access to monthly or yearly ED data would additionally allow segmented regression analysis, both to characterize whether the post-legalization increase occurred gradually or abruptly, and to assess whether the distribution of pamphlets and public health notices corresponds with subsequent decreases in CHS-related ED visits.

Local and state public health departments may also benefit from partnering with dispensaries, schools, and drug diversion programs to provide supplemental consumer education about CHS, with the goal of increasing awareness, reducing ED visits, and ultimately decreasing CHS incidence locally. The potential impact of these educational efforts could be assessed by tracking the distribution of CHS educational materials alongside local ED trends.

## Conclusion

This environmental scan addressed four research questions regarding the availability of CHS-related educational content and the local clinical burden of the condition in Kalamazoo, Michigan. Although this study could not establish a causal relationship between the availability of CHS-related educational content and the observed increase in ED encounters, it identified a clear and near-total absence of CHS-specific information across the consumer-facing resources evaluated, set against a substantial and growing clinical burden. Given the clinical and economic burden associated with delayed CHS recognition, these findings support the consideration of amending Michigan’s existing point-of-sale pamphlet requirement to include CHS-specific warning language. Similar disclosure gaps on retailer websites and public health resources merit consideration as well. Parallel efforts to strengthen clinician awareness, particularly in emergency and primary care settings, may improve CHS recognition, which is frequently misdiagnosed or underrecognized and can lead to repeated ED visits and diagnostic evaluations. This may help reduce diagnostic delay and associated costs, while closing the gap between the growing clinical burden of CHS and the limited consumer and provider education identified in this study.^27^

## Supporting information

Rubric

## List of abbreviations

CBD: Cannabidiol
CHS: Cannabinoid Hyperemesis Syndrome
ED: Emergency Department
MCRA: Michigan Cannabis Regulatory Agency
USD: United States dollar

## Declarations

### Ethics approval and consent to participate

This study was reviewed and approved by the Western Michigan University Homer Stryker M.D. School of Medicine Institutional Review Board (IRB # WMed-2026-1398). The requirement for informed consent was waived by the Institutional Review Board because of the retrospective nature of the study and the use of existing data.

### Consent for publication

Not applicable.

### Availability of data and materials

The dataset supporting the conclusions of this article is available in the Figshare repository at https://doi.org/10.6084/m9.figshare.33755530.

### Competing interests

The authors declare that they have no competing interests.

### Funding

This research was supported by institutional resources from Western Michigan University Homer Stryker M.D. School of Medicine.

### Authors’ contributions

ERL: conceptualization, data curation, methodology, project administration, writing-original draft, and review and editing. NB: formal analysis, investigation, and validation. LAB: investigation, validation, and writing-original draft. MEE: formal analysis, methodology, writing-original draft. DSR: writing-original draft. PVS: project administration, writing-review and editing. writing-original draft. HRW: visualization, writing-original draft. HZ: writing-original draft. JDM: supervision and writing-review and editing.

## Acknowledgements

The authors gratefully acknowledge Heidi Baker, R.N., Morgan Brown, and Theresa McGoff, M.B.A., R.N., of the Data Analytics Services Unit; Philip Kroth, M.D., of the Department of Biomedical Informatics; and Maureen Owens, M.M., and Madeline Rosenberger, B.S., of the Human Research Protection Program for their support and advice in this work.

## Declaration of Generative AI

During the preparation of this work, the authors used Elicit to brainstorm topic ideas, and AJE Curie and Paperpal to improve readability and language of the manuscript. After using these tools, the authors reviewed and edited the content as needed and assumed full responsibility for the content of the publication.

## Notes

### Competing Interest Statement

The authors have declared no competing interest.

### Summary of Updates

The ORCID added for co-authors Nicholas Bronson and Mary Engle. Correct in date format in methods section and Table 1 supplement.

## References

1. Allen JH, de Moore GM, Heddle R, Twartz JC. Cannabinoid hyperemesis: cyclical hyperemesis in association with chronic cannabis abuse. Gut. 2004;53(11):1566–70. doi: 10.1136/gut.2003.036350.

2. Michigan Legislature. Michigan Medical Marihuana Act: Initiated Law 1 of 2008, MCL 333.26421–333.26430. Lansing, MI: Michigan Legislature; 2008. [cited 2026 Sept 12]. Available from: https://legislature.mi.gov/Laws/MCL?objectName=MCL-333-26427.

3. Michigan Department of Licensing and Regulatory Affairs. Adult-use marijuana added to LARA’s renamed Bureau of Marijuana Regulation. Lansing, MI: State of Michigan; 2018 Dec 6. [cited 2026 Sept 12] Available from: https://www.michigan.gov/cra/spotlight/adult-use-marijuana-added-to-laras-renamed-bureau-of-marijuana-regulation.

4. Marijuana Policy Project. Michigan voters approve initiative to legalize and regulate marijuana for adult use [Internet]. Washington, DC: Marijuana Policy Project; 2018 Nov 6 [cited 2026 Sep 14]. Available from: https://www.mpp.org/news/press/michigan-voters-approve-initiative-to-legalize-and-regulate-marijuana-for-adult-use.

5. University of Michigan Injury Prevention Center. Impact of recreational cannabis legalization in Michigan: a baseline report. Ann Arbor (MI): University of Michigan; 2020 May. [cited 2026 Sept 12]. Available from: https://injurycenter.umich.edu/wp-content/uploads/2025/07/2020_IPC_Cannabis_Report.pdf.

6. City of Los Angeles Department of Cannabis Regulation. 2024 US cannabis sales [Internet]. Los Angeles, CA: City of Los Angeles; 2025 Apr 17 [cited 2026 Sep 14]. Available from: https://cannabis.lacity.gov/articles/2024-us-cannabis-sales.

7. U.S. Census Bureau. QuickFacts: Kalamazoo city, Michigan [Internet]. Washington, DC: U.S. Census Bureau; 2026 [cited 2026 Sep 14]. Available from: https://www.census.gov/quickfacts/fact/table/kalamazoocitymichigan/HSD310223.

8. Breuning M, Nie J, Schlesinger M, Smith L, Lavie-Ajayi M, Babinski S, Law S. The persistence of stigma and other challenges experienced by medical cannabis users: a thematic synthesis of qualitative studies. Med Cannabis Cannabinoids. 2026;9(1):115–149. doi: 10.1159/000551584.

9. Lamonica AK, Boeri M, Anderson T. Gaps in medical marijuana policy implementation: real-time perspectives from marijuana dispensary entrepreneurs, health care professionals and medical marijuana patients. Drugs Educ Prev Policy. 2016;23(5):422–434. doi:10.3109/09687637.2016.1150963.

10. Popova L, Massey ZB, Giordano NA. Warning labels as a public health intervention: effects and challenges for tobacco, cannabis, and opioid medications. Annu Rev Public Health. 2024;45(1):425–442. doi: 10.1146/annurev-publhealth-060922-042254.

11. Kalamazoo County Health & Community Services. Health & Community Services [Internet]. Kalamazoo (MI): Kalamazoo County; [cited 2026 Jul 15]. Available from: https://www.kalcounty.gov/236/Health-Community-Services.

12. Michigan Department of Health and Human Services. Michigan Department of Health and Human Services [Internet]. Lansing (MI): State of Michigan; [cited 2026 Jul 15]. Available from: https://www.michigan.gov/mdhhs.

13. Michigan Cannabis Regulatory Agency. Accela Citizen Access: cannabis licensing and regulatory records [Internet]. Lansing (MI): Michigan Department of Licensing and Regulatory Affairs; [cited 2026 Jul 15]. Available from: https://aca-prod.accela.com/mimm/Default.aspx.

14. Lorbeer E. Raw data. A growing burden, a persistent gap: cannabinoid hyperemesis syndrome education and emergency department trends in Michigan’s legal cannabis market [dataset]. Figshare; 2026. doi:10.6084/m9.figshare.33755530.

15. Medical Marihuana Facilities Licensing Act, 2016 PA 281, MCL 333.27206. Lansing (MI): Michigan Legislature; 2016. [cited 2026 Sept 12]. Available from: https://www.legislature.mi.gov/Laws/MCL?objectName=mcl-333-27206.

16. City of Kalamazoo. Chapter 20B: Marihuana commercial businesses, Article III: Specific marihuana business requirements, §20B-19: Provisioning centers and retailers. Kalamazoo (MI): City of Kalamazoo; 2022.

17. Michigan House Fiscal Agency. House Bills 4126 and 4127 as enacted: health warnings on marijuana products. Lansing (MI): Michigan Legislature; 2020 Apr 13. [cited 2026 Sept 12]. Available from https://www.legislature.mi.gov/documents/2019-2020/billanalysis/house/pdf/2019-HLA-4126-4CBB73F5.pdf.

18. The White House. Increasing medical marijuana and cannabidiol research. Executive Order No. 14370. Washington (DC): The White House; 2025 Dec 18. [cited 2026 Sept 12]. Available from: https://www.whitehouse.gov/presidential-actions/2025/12/increasing-medical-marijuana-and-cannabidiol-research/.

19. Johns T, Lawrence E. Evaluation and Treatment of Nausea and Vomiting in Adults. Am Fam Physician. 2024;109(5):417–425.

20. McFee RB. Cannabinoid Hyperemesis Syndrome (CHS) - an emerging gastrointestinal disorder and clinical challenge. Dis Mon. 2024;70(12):101832. doi: 10.1016/j.disamonth.2024.

21. Sandhu G, Smith S, Stephenson K, Jaeger V, John R, Shaver C, Johnson C. Prevalence of cannabinoid hyperemesis syndrome and its financial burden on the health care industry. Proc (Bayl Univ Med Cent). 2021;34(6):654–657. doi: 10.1080/08998280.2021.1937874.

22. Marshall A, Fai C, Han J, Yule AM, Jangi S. Rising inpatient utilization and costs of cannabis hyperemesis syndrome hospitalizations in Massachusetts after cannabis legalization. J Clin Gastroenterol. 2024;58(3):247–252. doi:10.1097/MCG.0000000000001857.

23. King DD, Gill CJ, Cadieux CS, Singh N. The role of stigma in cannabis use disclosure: an exploratory study. Harm Reduct J. 2024;21(1):21. doi: 10.1186/s12954-024-00929-8.

24. Meek CJ, Ranney LM, Clark SA, Jarman KL, Callanan R, Kowitt SD. Regulatory landscape of cannabis warning labels in US states with legal retail nonmedical cannabis, 2024. Am J Public Health. 2024;114(S8):S681–S684. doi: 10.2105/AJPH.2024.307722.

25. High Profile Cannabis. High Profile intro guide for the budding connoisseur: cannabis explained [Internet]. 2024 Jun [cited 2026 July 15] Available from: https://highprofilecannabis.com/wp-content/uploads/2026/06/High-Profile_Customer-Info-Booklet_CT_SC_2026_Digital-1v2.pdf.

26. Colorado General Assembly, Legislative Council Staff. State taxes on marijuana [Internet]. Denver, CO: Colorado General Assembly; 2014 Oct 31 [cited 2026 Sep 14]. Available from: https://content.leg.colorado.gov/publications/state-taxes-marijuana-2014.

27. Swartz JA, Franceschini D. Cannabinoid Hyperemesis Syndrome, 2016 to 2022. JAMA Netw Open. 2025 Nov;8(11):e2545310. doi: 10.1001/jamanetworkopen.2025.45310.

