## Supplementary material for "A Growing Burden, a Persistent Gap: Cannabinoid Hyperemesis Syndrome Education and Emergency Department Trends in Michigan’s Legal Cannabis Market": Rubric

**Supplemental Material 1**

**CHS Consumer Information Environmental Scan**

**Instructions: Coding Rubric & Data Collection Instrument**

Version 1 (2026)

**Purpose**

This instrument supports an environmental scan on cannabinoid hyperemesis syndrome (CHS). It is used to characterize the current landscape of consumer-facing CHS information across (1) licensed cannabis provisioning center websites in the city of Kalamazoo, Michigan, and (2) state public health department websites. Two reviewers should independently code each site, and discrepancies are resolved by discussion and recorded in the note column.

**Instructions for coders**

1. Navigate to Google Search, https://www.google.com, and perform a Google search on the name of the provisioning center or health department.
2. For each entry, record the site name, domain, and date accessed, even if no CHS content is found.
3. Code each of the five dimensions below using the categories defined in the rubric. Use the exact category of wording provided in the rubric to ensure that the responses can be tallied consistently.
4. If a site has no website or a website exists but contains no cannabinoid hyperemesis-health content, code Presence as "No mention" and leave the remaining dimensions blank.
5. For the provisioning center websites use Google search operators to locate relevant subpages for each identified domain: site:[domain] CHS OR "cannabinoid hyperemesis" OR “cannabis hyperemesis”
6. For the websites of the county public health department and state health department, use Google search operators: site:[domain] "cannabinoid hyperemesis" OR “cannabis hyperemesis”
7. Use the Notes column to document paywalled or login-gated content, inaccessible pages, and other issues that may affect the study or require discussion.

**Coding Rubric**

| **Dimension** | **Category** | **Definition** |
| --- | --- | --- |
| Presence | No mention | CHS is not mentioned |
|  | Mentioned by name | CHS is explicitly named |
| Depth | None | No CHS information |
|  | Warning Only | Brief earning (e.g., “may cause…”) |
|  | Moderate | Signs or symptoms listed |
|  | Substantial | Cessation guidance, when to seek medical help, or dedicated page |
| Web Page Placement | Not present | No CHS content |
|  | Buried | Found only through a Google search |
|  | Prominent | Accessible through top-level navigation |
| Audience | Consumer | Information intended for the public |
|  | Healthcare providers | Information intended for clinicians |
|  | Both | Intended for both audiences |
| Search String |  | Google search string used to identify CHS content |

**Data Collection Instrument**

One row per site (provisioning center or health department). Extend rows as needed.

| **Site Name** | **Domain** | **Date Accessed** | **Presence** | **Depth** | **Placement** | **Audience** | **Search String** |
| --- | --- | --- | --- | --- | --- | --- | --- |

Attribution: CHS consumer information environmental scan. Coding rubric and data collection instrument. Created by Elizabeth R. Lorbeer, EdM, MLS, Western Michigan University Homer Stryker M.D. School of Medicine, Kalamazoo, MI, USA
